# High household transmission of SARS-CoV-2 in the United States: living density, viral load, and disproportionate impact on communities of color

**DOI:** 10.1101/2021.03.10.21253173

**Authors:** Carla Cerami, Tyler Rapp, Feng-Chang Lin, Kathleen Tompkins, Christopher Basham, Meredith S. Muller, Maureen Whittelsey, Haoming Zhang, Srijana B. Chhetri, Judy Smith, Christy Litel, Kelly Lin, Mehal Churiwal, Salman Khan, Faith Claman, Rebecca Rubinstein, Katie Mollan, David Wohl, Lakshmanane Premkumar, Jonathan J. Juliano, Jessica T. Lin

## Abstract

**Background:** Few prospective studies of SARS-CoV-2 transmission within households have been reported from the United States, where COVID-19 cases are the highest in the world and the pandemic has had disproportionate impact on communities of color.

**Methods and Findings:** This is a prospective observational study. Between April-October 2020, the UNC CO-HOST study enrolled 102 COVID-positive persons and 213 of their household members across the Piedmont region of North Carolina, including 45% who identified as Hispanic/Latinx or non-white. Households were enrolled a median of 6 days from onset of symptoms in the index case. Secondary cases within the household were detected either by PCR of a nasopharyngeal (NP) swab on study day 1 and weekly nasal swabs (days 7, 14, 21) thereafter, or based on seroconversion by day 28. After excluding household contacts exposed at the same time as the index case, the secondary attack rate (SAR) among susceptible household contacts was 60% (106/176, 95% CI 53%-67%). The majority of secondary cases were already infected at study enrollment (73/106), while 33 were observed during study follow-up. Despite the potential for continuous exposure and sequential transmission over time, 93% (84/90, 95% CI 86%-97%) of PCR-positive secondary cases were detected within 14 days of symptom onset in the index case, while 83% were detected within 10 days. Index cases with high NP viral load (>10^6 viral copies/ul) at enrollment were more likely to transmit virus to household contacts during the study (OR 4.9, 95% CI 1.3-18 p=0.02). Furthermore, NP viral load was correlated within families (ICC=0.44, 95% CI 0.26-0.60), meaning persons in the same household were more likely to have similar viral loads, suggesting an inoculum effect. High household living density was associated with a higher risk of secondary household transmission (OR 5.8, 95% CI 1.3-55) for households with >3 persons occupying <6 rooms (SAR=91%, 95% CI 71-98%). Index cases who self-identified as Hispanic/Latinx or non-white were more likely to experience a high living density and transmit virus to a household member, translating into an SAR in minority households of 70%, versus 52% in white households (p=0.05).

**Conclusions:** SARS-CoV-2 transmits early and often among household members. Risk for spread and subsequent disease is elevated in high-inoculum households with limited living space. Very high infection rates due to household crowding likely contribute to the increased incidence of SARS-CoV-2 infection and morbidity observed among racial and ethnic minorities in the US. Quarantine for 14 days from symptom onset of the first case in the household is appropriate to prevent onward transmission from the household. Ultimately, primary prevention through equitable distribution of effective vaccines is of paramount importance.

**AUTHORS SUMMARY:** *Why was this study done?:*

- Understanding the secondary attack rate and the timing of transmission of SARS-CoV-2 within households is important to determine the role of household transmission in the larger pandemic and to guide public health policies about quarantine.
- Prospective studies looking at the determinants of household transmission are sparse, particularly studies including substantial racial and ethnic minorities in the United States and studies with adequate follow-up to detect sequential transmission events.
- Identifying individuals at high risk of transmitting and acquiring SARS-CoV-2 will inform strategies for reducing transmission in the household, or reducing disease in those exposed.

*What did the researchers do and find?:*

- Between April-November 2020, the UNC CO-HOST study enrolled 102 households across the Piedmont region of North Carolina, including 45% with an index case who identified as racial or ethnic minorities.
- Overall secondary attack rate was 60% with two-thirds of cases already infected at study enrollment.
- Despite the potential for sequential transmission in the household, the majority of secondary cases were detected within 10 days of symptom onset of the index case.
- Viral loads were correlated within families, suggesting an inoculum effect.
- High viral load in the index case was associated with a greater likelihood of household transmission.
- Spouses/partners of the COVID-positive index case and household members with obesity were at higher risk of becoming infected.
- High household living density contributed to an increased risk of household transmission.
- Racial/ethnic minorities had an increased risk of acquiring SARS-CoV-2 in their households in comparison to members of the majority (white) racial group.

*What do these findings mean?:*

- Household transmission often occurs quickly after a household member is infected.
- High viral load increases the risk of transmission.
- High viral load cases cluster within households - suggesting high viral inoculum in the index case may put the whole household at risk for more severe disease.
- Increased household density may promote transmission within racial and ethnic minority households.
- Early at-home point-of-care testing, and ultimately vaccination, is necessary to effectively decrease household transmission.

## INTRODUCTION

Since the onset of the COVID-19 pandemic, households have been a well-recognized setting for SARS-CoV-2 transmission. Proximity and ventilation, important determinants of person-to-person transmission [1], are difficult to control in shared living spaces. For those infected and isolating at home, following guidelines to sleep in a separate bedroom, use a separate bathroom, use masks, and not share items such as dishes, towels, and bedding [2] may be difficult in families with young children and/or small living spaces; especially once more than one household member is infected. Furthermore, since infectiousness and viral transmission peaks just before the onset of symptoms [3–5], household spread can occur before anyone is aware of a potential infection, as most Americans do not wear masks at home or in what they define as their family bubble.

Secondary household attack rates reported from China and other Asian countries early in the pandemic ranged from 10-15% [6]. This relatively low attack rate is at odds with anecdotal experience in the United States, where the virus has spread unchecked. While several meta-analyses have evaluated household transmission rates, all have incorporated both retrospective and prospective analyses. Prospective testing of household contacts regardless of symptoms status is required to estimate the true secondary attack rate (SAR). Yet only two such studies in the US have been reported. These two studies, following a total of 159 households in Utah, Wisconsin, and Tennessee, have started to paint a picture of much higher SARs in US households (29 and 53%) [7,8]. Yet, representation of racial and ethnic diversity was limited (around 25% of households), and testing was limited to 7 and 14 days of follow-up, which may not capture secondary cases that result from sequential transmission within households. Given the disproportionate impact of the COVID-19 epidemic on communities of color, measuring secondary household attack rates in vulnerable communities is important for shaping preventive and testing strategies, modeling spread, targeting high-risk populations, and assessing the length of time households should quarantine.

The UNC CO-HOST (COVID-19 Household Transmission Study) is the largest single-site observational household cohort in the US thus far and the most ethnically and racially diverse. Covering both suburban and rural areas of North Carolina, the study recruited from a testing center providing results within 24-hours that allowed for timely recruitment. Weekly sampling for quantitative viral loads combined with antibody testing at one month provided an extended period to evaluate transmission relative to other studies. During the time of this study, April to November 2020, the spike protein D614G variant was already fully penetrant in North Carolina [9]. The specific objective of this study was to measure the secondary attack rate in a setting where infected individuals were asked to quarantine at home and given standard guidance. Household and individual demographics as well as daily symptoms and weekly viral loads were collected to identify risk factors and timing of household transmission.

## METHODS

### Study Design

The CO-HOST Study evaluated SARS-CoV-2 transmission in the household of individuals who tested positive and quarantined at home. Here we describe the pre-planned primary analysis of the secondary attack rate and risk factors associated with SARS-CoV-2 transmission in the household setting in the southern United States. Study follow-up started in April 2020 and ended in November 2020.

### Ethics, standards and informed consent

The study was approved by the Institutional Review Board at the University of North Carolina and is registered at clinicaltrials.gov (NCT04445233). All participants (or their parents/guardians) gave written, informed consent. Minors over the ages of 7 provided assent.

### Study setting

Index cases were recruited after testing at the Respiratory Diagnostic Center at the University of North Carolina School of Medicine [10]. Participants were visited between 3-4 times at their private homes using a mobile unit van and returned to the Respiratory Diagnostic Center for the final study visit.

### Recruitment, screening and enrollment

Inclusion criteria for the index cases included any patient 18 years of age or older with a positive qualitative nasopharyngeal (NP) swab for SARS-CoV-2 obtained at UNC Hospitals, willingness to self-isolate at home for a 14-day period, willingness to participate in all required study activities for the entire 28-day duration of the study, living with at least one household contact who was also willing to consent to study follow-up, and living within reasonable driving distance (<1 hour) suitable for home visits by the study team. Inclusion criteria for household contacts of index patients included age greater than 1 year, and currently living in the same home as the index case without plans to leave to live elsewhere through the end of the 28-day study.

Pre-screening was conducted by telephone when qualifying results of the NP swab were available. During the telephone pre-screening, exclusion criteria were reviewed with the patient and the study procedures were reviewed with potential study participants.

The overall study design is depicted in **Figure S1**. After consenting, all participants were visited at their homes on Day 1 by a mobile clinical team. NP and nasal mid-turbinate (NMT) swabs were collected for analysis by PCR for SARS-CoV-2 and blood samples were collected for serology by both a rapid antibody test and an enzyme-linked immunosorbent assay (ELISA). Index cases and household contacts completed baseline questionnaires that included basic demographic and household information, abbreviated medical history, symptoms, recent travel history, and exposure to confirmed COVID-positive cases. All participants received instruction on how to perform a self-collected NMT swab. For nasal sampling, participants were instructed to insert the swab about 1-2 inches into one nostril, then swirl 5-8 times while slowly withdrawing the swab and placing it into the collection tube. In the case of participants under 7 years of age, parents or guardians were instructed how to perform the swabbing for their children.

All participants received a daily symptom questionnaire via email. Index cases and COVID-positive household contacts received the questionnaire daily until no symptoms were reported for two consecutive days. Other household contacts received the questionnaire daily for 21 days to monitor for symptoms that might indicate new COVID-19 infection.

On Days 7, 14 and 21, a study staff member conducted home visits for sample collection pickup. The staff member left a nasal swab on the doorstep for each participant and waited outside until everyone had completed the nasal swabs. At the final study visit on Day 28 participants were asked about COVID-related care-seeking and testing and underwent venipuncture for analysis of anti-SARS-CoV-2 antibodies by a rapid antibody test and by ELISA.

All samples collected during the study were placed into a cooler on ice immediately after collection and transported to a BSL2+ laboratory within 2 hours. If a study participant was hospitalized or left the household for other reasons, they were still followed until Day 28 to record outcomes, but sample collection was suspended.

### Laboratory analyses

#### qRT-PCR SARS-CoV-2 viral quantification

Nasopharyngeal and nasal swab samples were tested using a CDC RT-qPCR protocol authorized for emergency use that consists of three unique assays: two targeting regions of the virus’ nucleocapsid gene (N1, N2) and one targeting human RNase P gene (RP) (Catalog # 2019-nCoVEUA-01, Integrated DNA Technologies) [11]. Details of assay implementation and calculation of the limit of detection are described elsewhere [12]. Briefly, samples were designated positive if all three PCRs were positive (N1 and N2 for virus, RP for adequate sampling). The viral load of each sample, in copies/uL, was extrapolated from standard curves generated for each viral assay (N1 and N2) using serial dilutions of the nCoVPC plasmid control (2 to 100,000 viral RNA copies/uL). The average copies/uL between the N1 and N2 assays was used as the final quantitative viral load. Probit analysis yielded a limit of detection (LOD) for the N1 and N2 assays of 9 and 13 copies/uL, respectively. Thus, the average LOD between the two assays, 11 copies/uL, was used as the cutoff for sample positivity. Based on the sample collection and RNA extraction volumes as well as volume of template RNA used in the RT-qPCR (5uL), the reported viral load represents the number of viral RNA copies per 5 uL of VTM or Shield sample.

## Serology

### Rapid Test

The BioMedomics COVID-19 IgM/IgG Rapid Test is a point-of-care lateral flow immunoassay (LFIA) [13,14] that has been validated as a research tool [15]. Approximately 20 microliters of finger prick blood was obtained via a capillary sampler and dispensed on the sample port of the device. Two to three drops of buffer/developer solution were applied and results were read after 10 minutes by trained study staff. Positive, weak positive, and negative bands for IgM and IgG were recorded and a photograph was stored. A second reader reviewed the photographs blinded to the field results and consensus was reached on discrepant readings.

### Immunoassay to detect antibodies against the receptor binding domain (RBD) of the spike protein

Plasma samples were heat inactivated at 56°C for 30 minutes, then total Ig binding to the receptor binding domain (RBD) of the SARS-CoV-2 spike protein was measured using a previously described enzyme-linked immunosorbent (ELISA) assay [16,17]. Briefly, biotinylated recombinant antigen produced in mammalian cells consisting of SARS-2 Spike RBD is captured on a 96-well ELISA plate coated with streptavidin. The serum sample at 1:40 dilution is incubated with the RBD-captured wells, and bound antigen detected using HRP conjugated anti-goat total (IgG, IgM and IgA) antibody on a microplate reader. This in-house ELISA was previously evaluated on a large panel of well characterized samples and shown to have high sensitivity and specificity for detecting SARS-CoV-2 infection [16,17].

### D614G genotyping

A real-time PCR assay targeting a 107 bp region encompassing the D614G mutation in the SARS-CoV-2 spike protein receptor binding domain associated with increased viral load [18] was designed to evaluate the prevalence of 614G mutants in our study cohort. 5ul of RNA was reverse transcribed using the Invitrogen SuperScript III First-Strand Synthesis System for RT-PCR kit (Thermofisher Scientific). 2.5ul cDNA was then placed in 22.5uL of qPCR master mix with Roche FastStart Universal Probe Master (ROX) along with primers and probes listed in **Table S1**. Positive control plasmids for mutant (MT) and wild-type (WT) sequences were synthesized by Genewiz (inserts listed in **Table S1**) and used to set the appropriate Ct threshold for positivity in each run. Samples were considered WT if detected only by WT probe; MT if detected only by MT probe or if detected by both MT and WT probes with MT Ct >3 cycles lower than WT Ct; or mixed (containing both WT and MT virus) if detected by both with Ct difference of <3 cycles.

### Sample size determination

This is a prospective observational study. The planned target enrollment was 200 households. The study was stopped prior to reaching this target due to funding considerations.

### Study objectives and outcomes

The primary objective was to evaluate the secondary household attack rate among household members of persons quarantined in their home after testing positive for SARS-CoV-2.

The primary study endpoint was SARS-CoV-2 infection in the household contacts as determined by real-time PCR of nasopharyngeal or nasal swabs for SARS-CoV-2 at any of the timepoints or evidence of seroconversion during the study based on anti-SARS-CoV-2 antibody testing.

A secondary objective was to assess individual and household risk factors associated with SARS-CoV-2 transmission in the household.

### Data entry, handling, storage and security

After giving written consent, the participants were given a study identification number, which was used in all future datasets for participant anonymity. Collected data were entered in real-time using electronic Case Report Forms (eCRF) developed on a REDCap (Research Electronic Data Capture) database. Any data collected on paper format was entered by a study staff member and then checked by the study coordinator. Daily symptom diaries were entered directly into the REDCap database by the participants and were checked by study staff for completion and inconsistencies. Laboratory related data were extracted directly from laboratory equipment and uploaded to the database. The study was conducted in compliance with Good Clinical Practice.

### Statistical analysis

For each household, if multiple participants were SARS-CoV-2 positive at enrollment, we defined the index case as the person with the earliest onset of infection based on onset of symptoms and known date(s) of PCR test positivity. If this was ambiguous and to prevent bias, then baseline antibody positivity was also used as evidence of less recent infection. This resulted in index case reassignments in 11 households. Any study participant with evidence of prior infection (antibody-positive with negative PCR) at enrollment was excluded from the analysis (n=4).

We summarized demographic characteristics and underlying conditions of index cases and household contacts, as well as their household demographics. Baseline characteristics that are continuous variables were dichotomized (e.g. age, BMI) per standard conventions.

The secondary attack rate (SAR) among household contacts was calculated as the proportion of susceptible household contacts with laboratory-confirmed SARS-CoV-2 infection during the 28-day follow-up period. Household contacts who were COVID-positive at enrollment and reported the same COVID exposure outside the household as the index case were not considered in the at-risk population as susceptible contacts. As per above, secondary cases were defined as the remaining susceptible household contacts found positive for SARS-CoV-2 by PCR testing or with evidence of seroconversion during the study. Household contacts were excluded from the SAR analysis if they missed all follow-up study visits (n=6) or were symptomatic with negative PCR testing but missing antibody data at day 28 (n=1). Among those included in the analysis, the rate of missing data was low (<5%); thus, we did not impute missing data. A 95% CI for the SAR was constructed using the Wilson method for a single proportion. A logistic regression model with a random intercept to account for within-household variation was used to calculate the race/ethnicity-specific SAR.

In the primary SAR analysis, all secondary cases were presumed due to household transmission (not community-acquired). Sensitivity analyses were performed excluding secondary cases already infected at baseline or excluding secondary cases identified at day 14 or later that may have been acquired outside the household. The SAR for households was calculated as the proportion of households with at least one secondary case identified in the household during the 28-day follow-up.

We estimated the serial interval (in days) of symptom onset between sequential SARS-CoV-2 infections in the household, as well as the number of days between symptom onset of the index case and PCR positivity of secondary cases in the household.

We determined whether nasopharyngeal SARS-CoV-2 viral loads were correlated within households (whether persons in the same household were more likely to have similar NP viral loads) by the intraclass correlation coefficient (ICC), which compares within versus between households variation of baseline NP viral loads. For those participants who did not complete an NP swab on study day 1, we used a transformed NMT viral load to impute the missing NP value. The transformation formula was derived from a linear regression equation generated from >100 study participants with positive viral load from both NP and NMT swabs on study day 1 [12]. To determine whether NP viral load in index cases was associated with secondary cases in the same household, we dichotomized the NP viral load with a cutoff of 1×10^6 viral copies/ul and compared the proportion of transmission events.

Finally, we examined other potential risk factors for secondary transmissions within the household, including characteristics of index cases, household contacts, and their household environment. We presented the odds ratio (OR) and corresponding 95% CI for potential risk factors using logistic regression with a random intercept to account for within-household correlation. Household contacts were excluded from the risk factors analysis if they missed all follow-up study visits (n=3) unless they were already found to be infected at enrollment (n=3). To address potential misclassification, we excluded household contacts with negative PCR testing but missing antibody testing at day 28 (n=3).

Statistical analysis and preparation of figures were conducted using R 4.0.2 (R Core Team, Vienna, Austria), GraphPad Prism (GraphPad Software INC, CA 92037, USA), and ArcGIS (Esri, Redlands, California). All hypothesis tests were two-sided at a significance level of 0.05 with no adjustment for multiplicity.

## RESULTS

### Household enrollment and demographics

Between April 29 - October 16, 2020, the UNC CO-HOST study recruited and enrolled 102 households all of whom had at least one member with laboratory confirmed SARS-CoV-2 infection. Two households were excluded from analysis because all household contacts either had evidence of prior infection at baseline (antibody-positive with negative PCR test) or did not complete the baseline questionnaire. The remaining 100 households (median size = 3.5 persons) were enrolled a median of 6 (IQR 4-7) days after symptom onset of the designated index case. These households spanned 34 zip codes across the North Carolina Piedmont Region, North Carolina, USA (**Figure 1**).

**Figure 1.**
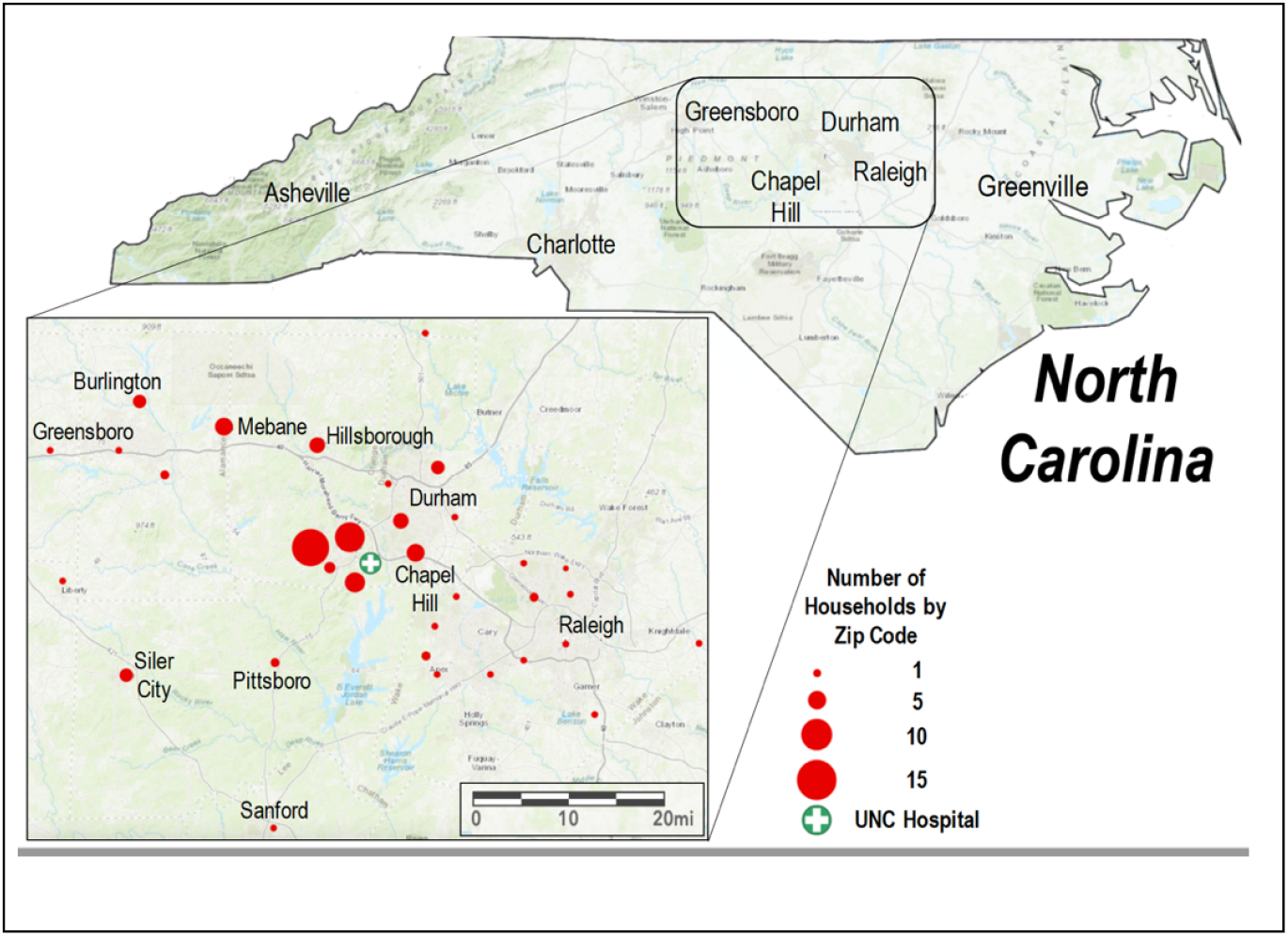
Geography of 100 households enrolled the UNC CO-HOST study.

Among the 100 participating households, the index case was reassigned in 11 households. Four household contacts were antibody-positive but PCR-negative at enrollment (indicating prior infection) and thus excluded from analysis. One household contact without antibody data at either day 1 or day 28 was also excluded. Baseline characteristics for the remaining 100 index cases and 204 household contacts (HCs) enrolled in the study are shown in **Table 1**.

**Table 1.**
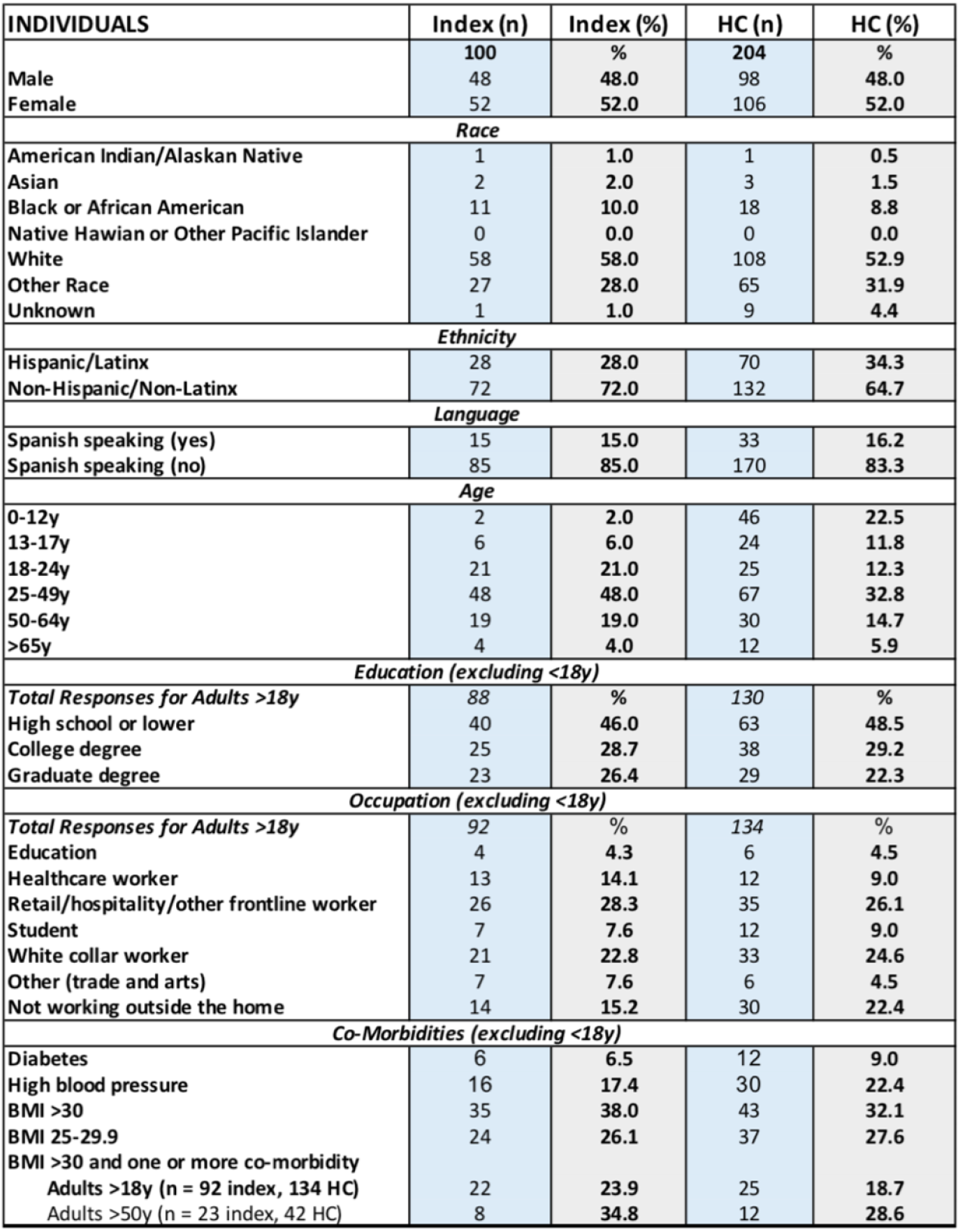
Demographics of study participants.

Among the 100 index cases, 48 were male, 52 were female, 92 were over 18 years of age and 42 reported non-white race-ethnicity. The index cases had a median viral load of 148,992 copies/ul (IQR 757-2,423,155 copies/ul) at the first study visit on nasopharyngeal (NP) swab. Among the 204 household contacts, 48% were male, 52% were female, 66% were over 18 years of age and 47% reported non-white race-ethnicity. Both the index cases and HCs had a similar percentage of adult participants with a Body Mass Index (BMI) over 30 kg/m2: 38% of index cases and 32% of household contacts, consistent with the prevalence of obesity in North Carolina (34%)[19]. A significant number of adult index cases (24%) and household contacts (19%) had both obesity and one other co-morbidity. Further description of the underlying conditions is shown in **Table S2**. Three index cases and three household contacts (all from different households) also enrolled in a treatment study in which they were randomized to receive either the oral drug EIDD-2801 (molnupiravir) or placebo (NCT04405570).

Household demographics are shown in **Table S3**. 27% of participating households were limited to two members, while 28% of households had 5 or more members. 63% were owner occupied single family homes and 42% lived in homes greater than 2,000 square feet. Households with a non-white index case were larger (median household size 4 versus 3, p=0.02) and also more likely to live in a home <2,000 square feet (76% versus 43%, p=0.003) compared to households with a white index case. This led to a higher “living density” for non-white households: 41% had >3 household members living in a home with fewer than 6 rooms, compared to 10% of white households (p<0.001). In 44% of households, at least one household member declined to be enrolled in the study.

### Secondary attack rate among household contacts

The overall secondary attack rate (SAR) among susceptible household contacts was 60% (106/176, 95% CI 53%-67%) (**Figure 2**). Of 100 households with 304 study participants (100 index cases and 204 HCs) included in the analysis, 99 households completed one month follow-up. One household of 6 withdrew shortly after enrollment. No households were lost to follow-up. Twenty-two of the household contacts tested positive at baseline for SARS-CoV-2, but were judged to have had the same environmental exposure to SARS-CoV-2 as the index cases (for example, both attended a cookout or other gathering where multiple individuals later tested COVID-positive). These contacts were considered to have a common exposure with the index case and were excluded from the transmission analysis, leaving 176 susceptible HCs.

**Figure 2.**
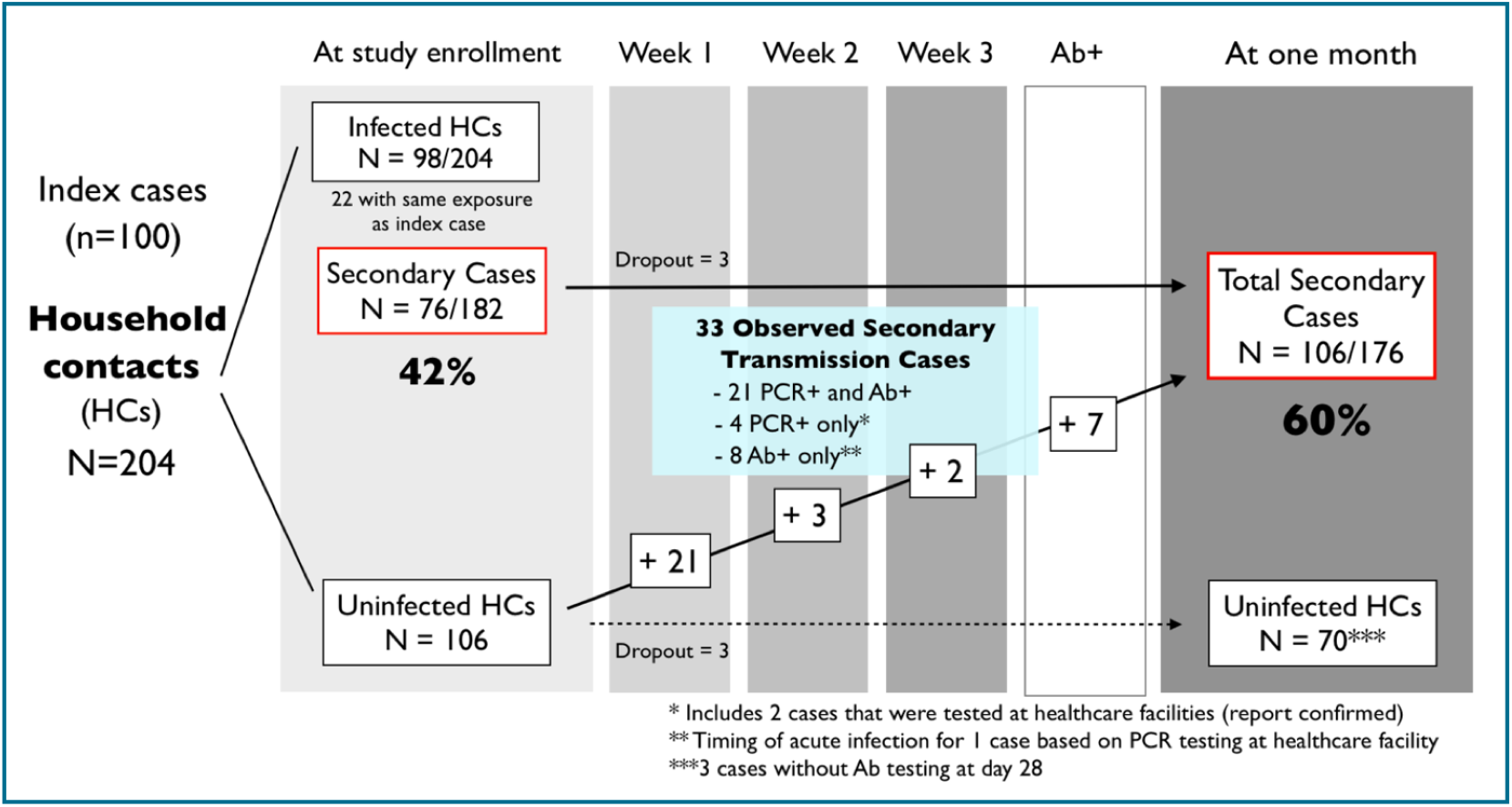
Secondary attack rate (SAR) among susceptible household contacts. Of 100 households included in the analysis, 99 completed one month follow-up. One household of 6 withdrew (3 infected at baseline). Among 182 susceptible household contacts, 42% (76/182) were already infected at the time of study enrollment and 33 additional secondary cases were observed during follow-up, resulting in an overall SAR of 60% (106/176,95% CI 53%-67%).

Secondary transmission cases were defined as household members who either tested positive for SARS-CoV-2 either by PCR or had evidence of seroconversion by day 28. Among the 176 susceptible household contacts, 73 were positive for SARS-CoV-2 at baseline (plus 3 that dropped out) and were classified as secondary cases. 33 additional secondary cases were observed during the study follow-up. Thus, 42% of HCs were already infected at the time of study enrollment, while the cumulative SAR was 60% (106/176, 95% CI 53%-67%). Among those infected at enrollment, 90% (64/71) reported having symptoms within the previous week, with a median duration of 5 days of symptoms at the time of enrollment.

Of the 33 secondary transmission cases that were observed during the study, 25 were identified by PCR testing and 8 were detected only because they seroconverted and were antibody positive at the day 28 visit. The majority (n=21) occurred in the first week after enrollment. Of the 5 cases detected by PCR after the first week of enrollment, 4 occurred in households of 5 or more, including 2 from the same household. Of the 33 secondary cases among household contacts who became infected with SARS-CoV-2 during the study, 27 (82%) experienced symptoms while 6 (18%) remained asymptomatic.

If restricting the SAR to a more conservative definition of only those secondary cases that were observed during the study (i.e. those who tested negative at baseline), the observed SAR was 32% (33/103). If removing late secondary cases that were identified at study day 14 or later, considering that these may have been acquired via later community exposure rather than household transmission, the early SAR ranged between 53-57% (depending on how the 7 cases identified only by antibody-positivity are distributed).

At the household level, assessing whether any secondary cases occurred within the household, SAR was even higher and skewed towards early transmission (**Figure 3**). Fifty three percent of susceptible households (49/92) contained at least one infected household member at enrollment besides the primary index case, rising to 70% (64/92) of households containing secondary cases one month later.

**Figure 3.**
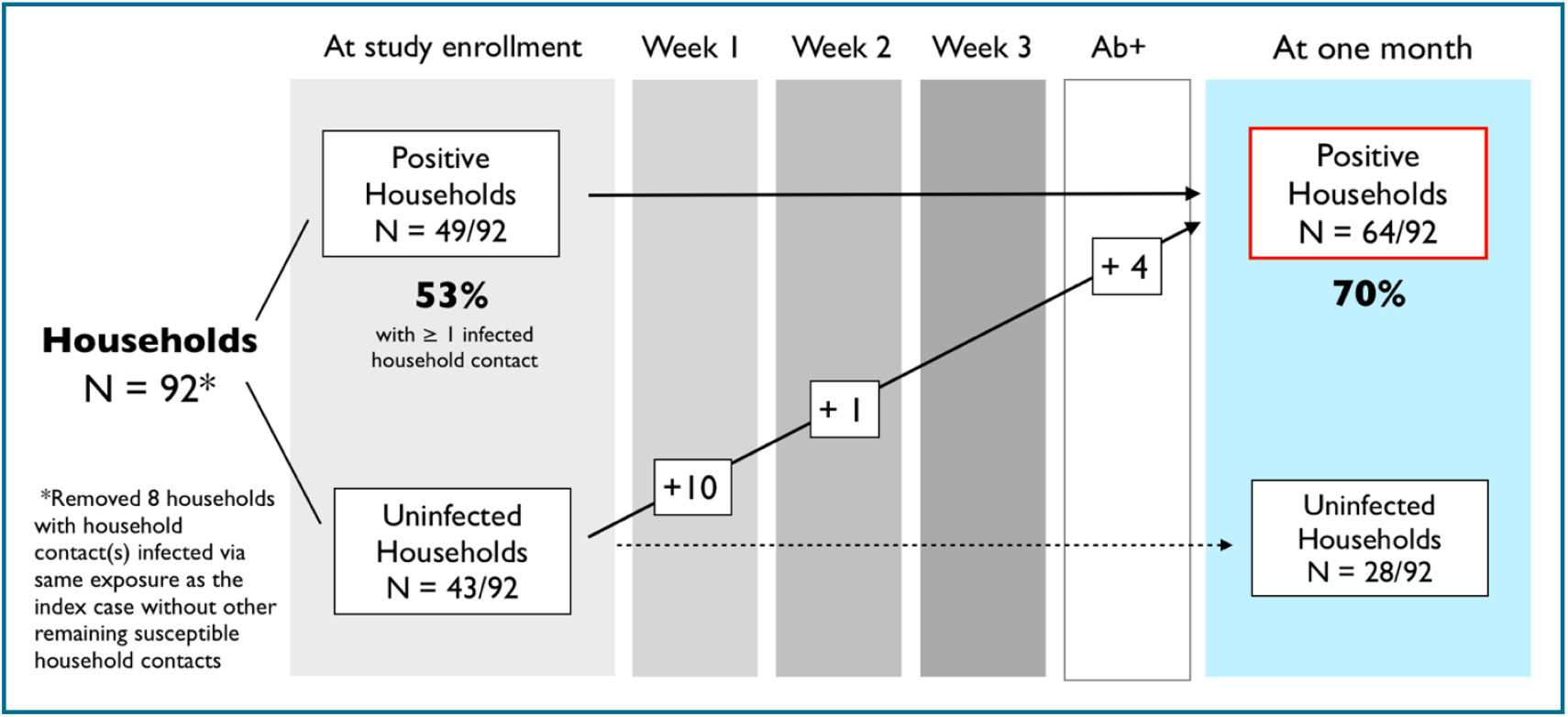
Secondary household attack rate. Of 92 households included in the analysis, 53% (49/92) contained infected household contacts at enrollment, with 15 more households sustaining transmission over the next 21 days, resulting in a secondary household attacak rate of 70% (64/92).

### Timing of secondary cases within the household

The serial interval for secondary cases in the household, based on onset of symptoms was a median of 3 days (IQR 1-6 days) after symptom onset in the index case and 2 days (IQR 1-4 days) from the most recent symptomatic case in the household. Because over two-thirds of secondary household cases (73/106 or 69%) were already infected at enrollment and 28% of households had multiple secondary cases, we regard these as imprecise estimates.

However, understanding when secondary cases became PCR-positive in relation to onset of symptoms in the index or other preceding case(s) is useful for informing guidelines for duration of quarantine [20]. Of the 89 PCR+ secondary cases for which the index case reported symptom duration, 84% (75/89) tested PCR-positive within 10 days of illness onset in the index case, while 94% (84/89) tested PCR-positive within 14 days. When also taking into account other subsequently infected household members besides the index case, 93% (83/89) of secondary cases tested PCR+ within 10 days of reported symptom onset of the most recent case in the same household while 99% (88/89) tested PCR-positive within 14 days. Thus, “resetting the clock” on a 14-day quarantine period based on subsequent COVID+ cases in the household would have achieved incremental benefit, isolating 4 more cases during the extended quarantine period. One of these was an asymptomatic infection with low viral load (402 copies/ul on NMT swab) found at study day 14, while the other 3 cases (2 from the same household) were symptomatic prior to their PCR diagnosis.

### Viral load within households and transmission

SARS-CoV-2 viral burden is correlated within households (**Figure 4**). When comparing the baseline nasopharyngeal viral load within versus between households, viral burden showed significant clustering within households (ICC=0.44, 95% CI 0.26-0.60, p<0.001). Differences in viral load are not attributable to D614G mutation in the viral spike protein that has been associated with increased viral load and infectivity [18], as the vast majority of isolates genotyped contained the mutation. Of 92 COVID-positive isolates (index cases and HCs) that were successfully genotyped from the first 90 households, 90/92 (98%) contained the 614G mutant, while only 2 were wild-type at this locus.

**Figure 4.**
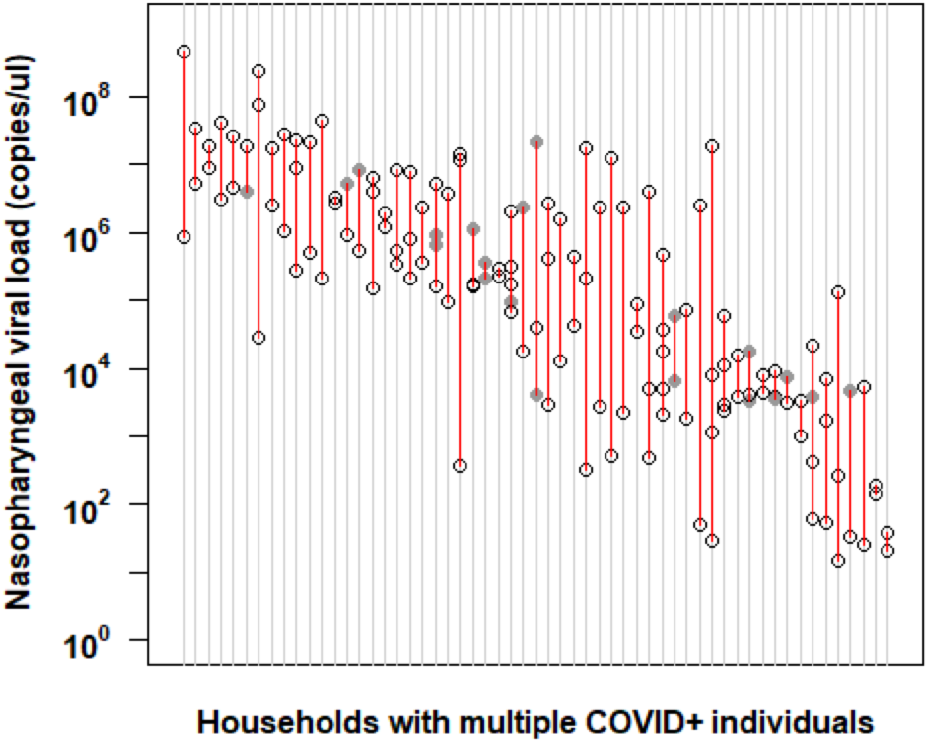
SARS-CoV-2 viral burden is correlated within families. The viral load obtained at enrollment from nasopharyngeal swabs in households with multiple COVID-positive household members are shown. Each vertical row in red depicts an individual household, with circles delineating the log viral load of each member within the household. Circles shaded in gray represent values derived from a nasal mid-turbinate swab if NP sampling was not performed. Households are depicted across the x-axis in order of decreasing viral load. Data drawn from 98 households and 184 participants. The intraclass correlation coefficient ICC = 0.44, 95% Cl (0.26, 0.60), p-value < 0.001.

Additionally, index cases with a high NP viral load (>10^6 viral copies/ul) at study enrollment were more likely to transmit virus to their household contacts during the study (OR 4.9, 95% CI 1.3-18 p=0.02). The median NP viral load among index cases was 1.4 log_10_ higher in households with new secondary cases detected during the study versus those with no transmission in the household (**Figure 5**). This difference was even greater when restricting the analysis to index cases who were not already antibody-positive, and thus more recently infected [15,16]. This association of index viral burden and transmission did not extend to secondary cases that were already present at study enrollment, likely due to a failure to capture the peak viral load of the index case in these households. Other characteristics of COVID disease status of the index case - including duration of symptoms and symptom severity - were not associated with secondary transmission in the household (**Table 2**). However, the 4 index cases that were hospitalized transmitted within the household before hospitalization.

**Table 2.** Potential risk factors for SARS-CoV-2 transmission from index cases.

| INDEX CASES | All Indexes<br>(n, %) | Transmitters<br>(n, %) | Non-transmitters<br>(n, %) | p-value |
| --- | --- | --- | --- | --- |
|  | 92 (100%) | 64 (70%) | 28 (30%) | - |
| *Age* |  |  |  |  |
| <18y | 8 (9%) | 5 (8%) | 3 (11%) | NS |
| 18-50y | 64 (70%) | 44 (69%) | 20 (71%) | NS |
| >50y | 20 (22%) | 15 (23%) | 5 (18%) | NS |
| *Sex* |  |  |  |  |
| Female | 49 (53%) | 31 (48%) | 18 (64%) | NS |
| Male | 43 (47%) | 33 (52%) | 10 (36%) | - |
| *Mask wearing prior to enrollment (missing n = 4)* |  |  |  |  |
| Mask wearing at home | 15 (17%) | 9 (15%) | 6 (22%) | NS |
| *Race/Ethnicity* |  |  |  |  |
| White, non-Hispanic | 51 (55%) | 30 (47%) | 21 (75%) | 0.02 |
| Black or African American | 10 (11%) | 8 (13%) | 2 (7%) | NS |
| Other, non-Hispanic | 5 (5%) | 5 (8%) | 0 (0%) | NS |
| Hispanic/Latinx | 26 (28%) | 21 (33%) | 5 (18%) | NS |
| *Symptom severity (missing n = 5)* |  |  |  |  |
| Mild | 25 (29%) | 17 (29%) | 8 (29%) | NS |
| Moderate/Severe | 58 (67%) | 38 (64%) | 20 (71%) | NS |
| Hospitalized | 4 (5%) | 4 (7%) | 0 (0%) | NS |
| *Duration of symptoms at enrollment (missing n = 8)* |  |  |  |  |
| Median (IQR) | 6 (4-7) | 6 (5-7) | 6 (4-7) | NS |
| *Antibody status at enrollment (missing n = 4)* |  |  |  |  |
| IgG-positive | 32 (36%) | 24 (39%) | 8 (30%) | NS |
| IgG-negative | 51 (58%) | 34 (56%) | 17 (63%) | NS |
| *Co-morbidities for adults >18y (missing n = 1 for diabetes, n = 5 for obesity)* |  |  |  |  |
| Diabetes | 6 (7%) | 6 (9%) | 0 (0%) | NS |
| Obesity, BMI >30 | 34 (39%) | 26 (43%) | 8 (30%) | NS |
| *Education for adults >18y (missing n = 12)* |  |  |  |  |
| High school or lower | 36 (45%) | 28 (51%) | 8 (32%) | NS |
| College degree | 23 (29%) | 15 (27%) | 8 (32%) | NS |
| Graduate degree | 21 (26%) | 12 (22%) | 9 (36%) | NS |
| *Occupation* |  |  |  |  |
| Healthcare worker | 13 (14%) | 5 (8%) | 8 (29%) | 0.01 |

p-values only reported if $\leq 0.10$ , otherwise noted as not significant (NS)

**Figure 5.**
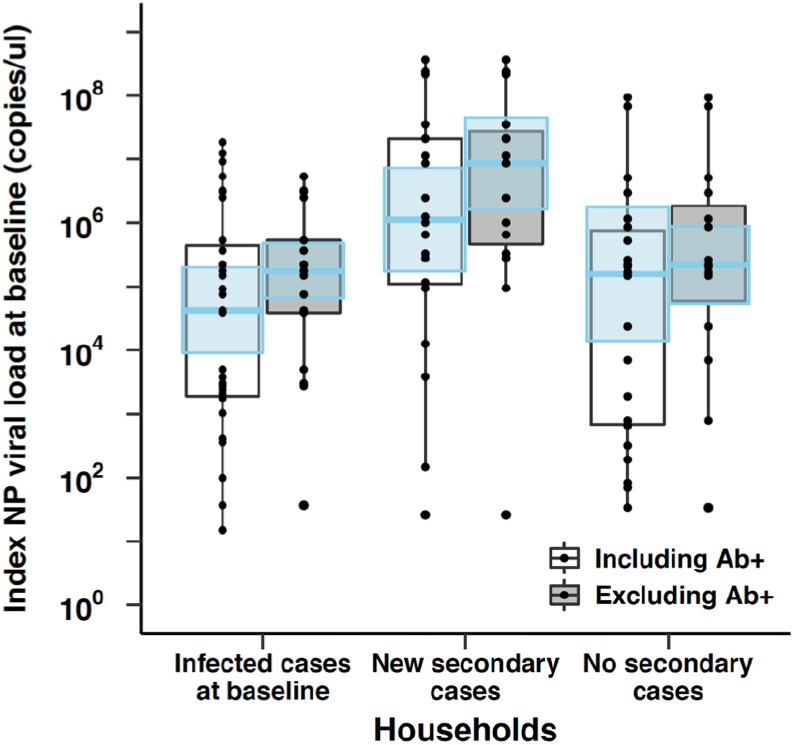
Association of index nasopharyngeal viral load and transmission in the household. Households with new secondary cases folllowing enrollment were more likely to have index cases with high nasopharyngeal viral load compared to households without secondary transmission. Index cases that were not antibody-positive at enrollment, as a marker of more recent infection, are depicted to the right in gray. Blue overlaid boxes depict 95% CIs.

### Other risk factors for household transmission

Non-white index cases were more likely to transmit virus within their household (**Table 2**), despite there being no difference in viral loads by race/ethnicity (data not shown). This translates to a SAR of 70% (95% CI 59%-79%) in households where the index case was non-white or Hispanic compared to 52% (95% CI 42%-62%) in white households (**Table 3**). Among other factors, this is likely attributable to household crowding. A higher living density, defined as greater than 3 household members living in a home with fewer than 6 rooms (excluding bathrooms and garage), was associated with a greater odds of infection (OR 5.9, 95% CI 1.3-27; SAR 91%, 95% CI 71%-98% in high living density households) (**Table 4**), and a greater proportion of non-white/Hispanic households met this definition of high living density (44%, 18/41) compared to white households (8%, 4/51) (p< 0.001). Healthcare workers were less likely to transmit virus within the household (OR 0.22 95% CI 0.05-0.85) (**Table 2**).

**Table 3.** Secondary attack rate by race/ethnicity of index case in the household.

| Race/Ethnicity | Susceptible<br>HCs | Secondary household transmission |  |  | SAR (95% CI) |
| --- | --- | --- | --- | --- | --- |
|  |  | at baseline | over 21 days | total* |  |
| All | 176 | 76 | 33 | 106* | 60% (53-67%) |
| White, non-Hispanic | 96 | 32 | 18 | 50 | 52% (42-62%) |
| Non-white | 80 | 41 | 15 | 56 | 70% (59-79%) |
| Black or African-American | 17 | 8 | 4 | 12 | 71% |
| Hispanic/Latinx | 56 | 33 | 10 | 40* | 71% |
| Other, non-Hispanic | 7 | 3 | 1 | 4 | 57% |

\*accounting for 3 dropouts from secondary cases infected at baseline

**Table 4.** Potential household-level risk factors for SARS-CoV-2 transmission.

| HOUSEHOLDS | All Households (n, %) | Infected (n, %) | Uninfected (n, %) | p-value |
| --- | --- | --- | --- | --- |
|  | 92 (100%) | 64 (70%) | 28 (30%) | - |
| *Household size* |  |  |  |  |
| Mean | 3.8 | 3.9 | 3.4 | NS |
| *Living space (missing n = 5)* |  |  |  |  |
| <2000 sq ft | 48 (55%) | 37 (62%) | 11 (41%) | NS |
| >2000 sq ft | 39 (45%) | 23 (38%) | 16 (59%) | 0.07 |
| *Number of rooms\** |  |  |  |  |
| 2 or fewer rooms | 10 (11%) | 7 (11%) | 3 (11%) | NS |
| 3-5 rooms | 40 (43%) | 32 (50%) | 8 (29%) | NS |
| 6 or more rooms | 42 (46%) | 25 (39%) | 17 (61%) | NS |
| *Living density* |  |  |  |  |
| >3 people and <6 rooms | 22 (24%) | 20 (31%) | 2 (7%) | 0.02 |
| *Home ownership (missing n = 4)* |  |  |  |  |
| Renting apartment | 10 (11%) | 8 (13%) | 2 (7%) | NS |
| Renting home | 25 (28%) | 19 (32%) | 6 (21%) | NS |
| Own home | 53 (60%) | 33 (55%) | 20 (71%) | NS |

\*Number of rooms includes bedrooms, kitchen, and common rooms, but not bathrooms or garage p-values only reported if $\leq 0.10$ , otherwise noted as not significant (NS)

Among suscep ble household contacts, partners of the index case and those with a BMI in the obesity range were at higher risk of acquiring infec on(OR 4.1, 95% CI 1.3-13 and OR 5.4, 95% CI 1.4-21, respec vely)(**Table 5**). While not reaching sta s cal significance, non-white household members and those who shared a bedroom with the index case appeared to have a higher risk of 419 infec on. Sharing a bathroom was associated with a higher risk of secondary infec on during study follow-up (p=0.01, data not shown). Children of the index case a lower risk of 422 infec on, but this did not reach sta s cal significance (OR 0.42, 95% CI -1.2).

**Table 5.**
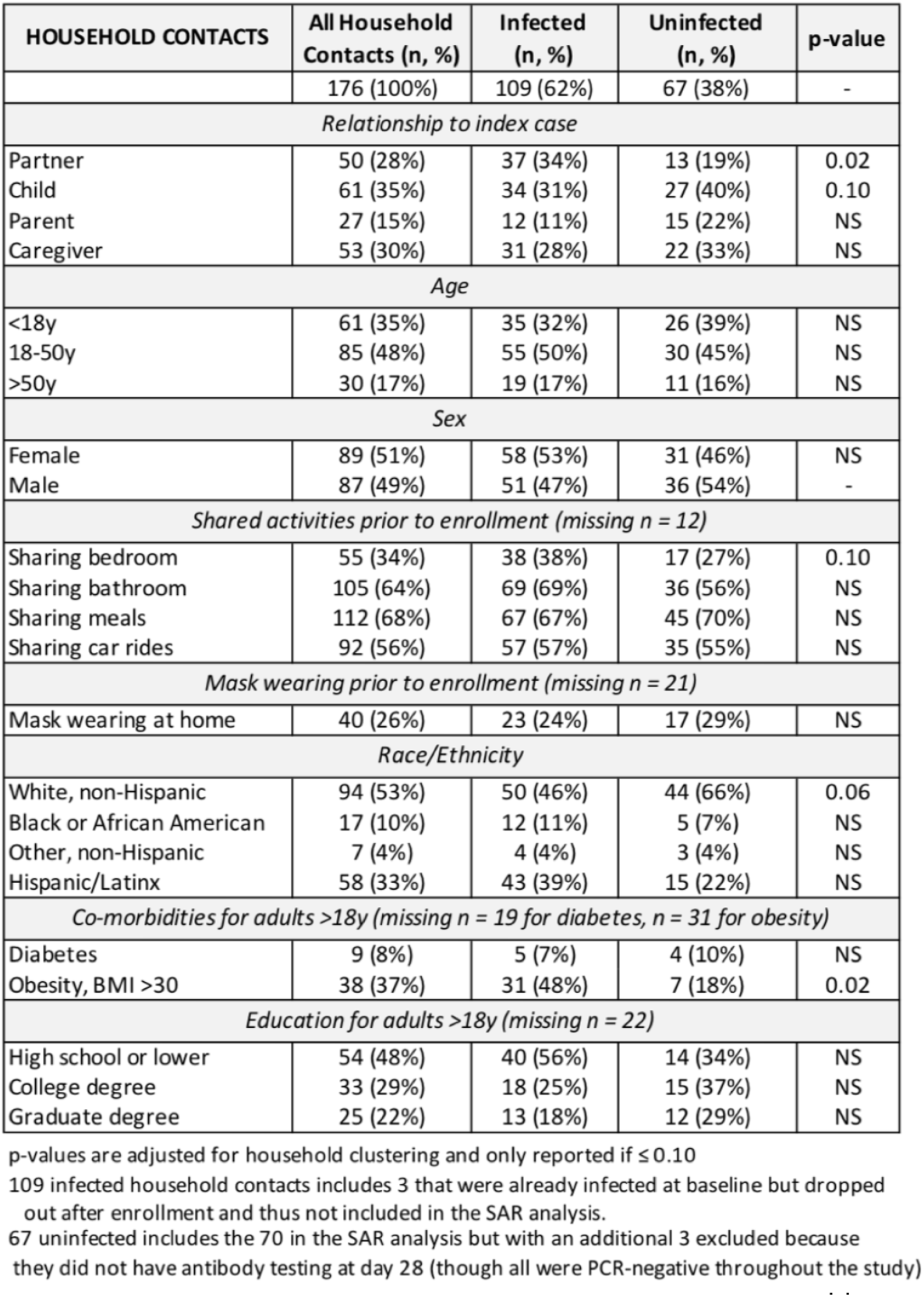
Potential risk factors for SARS-CoV-2 infection among household contacts.

| HOUSEHOLD CONTACTS | All Household Contacts (n, %) | Infected (n, %) | Uninfected (n, %) | p-value |
| --- | --- | --- | --- | --- |
|  | 176 (100%) | 109 (62%) | 67 (38%) | - |
| *Relationship to index case* |  |  |  |  |
| Partner | 50 (28%) | 37 (34%) | 13 (19%) | 0.02 |
| Child | 61 (35%) | 34 (31%) | 27 (40%) | 0.10 |
| Parent | 27 (15%) | 12 (11%) | 15 (22%) | NS |
| Caregiver | 53 (30%) | 31 (28%) | 22 (33%) | NS |
| *Age* |  |  |  |  |
| <18y | 61 (35%) | 35 (32%) | 26 (39%) | NS |
| 18-50y | 85 (48%) | 55 (50%) | 30 (45%) | NS |
| >50y | 30 (17%) | 19 (17%) | 11 (16%) | NS |
| *Sex* |  |  |  |  |
| Female | 89 (51%) | 58 (53%) | 31 (46%) | NS |
| Male | 87 (49%) | 51 (47%) | 36 (54%) | - |
| *Shared activities prior to enrollment (missing n = 12)* |  |  |  |  |
| Sharing bedroom | 55 (34%) | 38 (38%) | 17 (27%) | 0.10 |
| Sharing bathroom | 105 (64%) | 69 (69%) | 36 (56%) | NS |
| Sharing meals | 112 (68%) | 67 (67%) | 45 (70%) | NS |
| Sharing car rides | 92 (56%) | 57 (57%) | 35 (55%) | NS |
| *Mask wearing prior to enrollment (missing n = 21)* |  |  |  |  |
| Mask wearing at home | 40 (26%) | 23 (24%) | 17 (29%) | NS |
| *Race/Ethnicity* |  |  |  |  |
| White, non-Hispanic | 94 (53%) | 50 (46%) | 44 (66%) | 0.06 |
| Black or African American | 17 (10%) | 12 (11%) | 5 (7%) | NS |
| Other, non-Hispanic | 7 (4%) | 4 (4%) | 3 (4%) | NS |
| Hispanic/Latinx | 58 (33%) | 43 (39%) | 15 (22%) | NS |
| *Co-morbidities for adults >18y (missing n = 19 for diabetes, n = 31 for obesity)* |  |  |  |  |
| Diabetes | 9 (8%) | 5 (7%) | 4 (10%) | NS |
| Obesity, BMI >30 | 38 (37%) | 31 (48%) | 7 (18%) | 0.02 |
| *Education for adults >18y (missing n = 22)* |  |  |  |  |
| High school or lower | 54 (48%) | 40 (56%) | 14 (34%) | NS |
| College degree | 33 (29%) | 18 (25%) | 15 (37%) | NS |
| Graduate degree | 25 (22%) | 13 (18%) | 12 (29%) | NS |

p-values are adjusted for household clustering and only reported if $\leq 0.10$
109 infected household contacts includes 3 that were already infected at baseline but dropped out after enrollment and thus not included in the SAR analysis.
67 uninfected includes the 70 in the SAR analysis but with an additional 3 excluded because they did not have antibody testing at day 28 (though all were PCR-negative throughout the study)

## DISCUSSION

Household transmission is one of the main drivers of the SARS-CoV-2 pandemic. By incorporating timely recruitment of index cases, prospective sampling to 21 days regardless of symptom status, and diverse representation, we show that household transmission occurs in the majority of COVID-positive North Carolina households. The overall secondary attack rate in our sample was 60%, rising to 70% in minority households and 91% in households with higher living density. Importantly, we show not only that those infected with a high viral load are more likely to transmit virus to other members of the household, but that they seed other high-viral load infections, putting the entire household at higher risk for more severe illness [21]. Spread within the household happens quickly, often with one or more household members already infected by the time the first case in the household is diagnosed.

While the most complete meta-analysis of household transmission studies, published in December 2020, found a much lower overall household SAR of 16.6% (95% CI, 14.0%-19.3%), it noted significant heterogeneity between studies (ranging 4-45%) and combined both retrospective studies based on contact tracing data and prospective analyses, with the former comprising most of the studies [6]. As would be expected, studies with increased frequency of testing regardless of symptom status generally show higher infection rates [22]. In the US, a retrospective study in New York that included household testing offered regardless of symptom status reported a SAR of 38% [23], while two more recently published prospective studies following a total of 159 households in Utah and Wisconsin (58 households, SAR 29%)[7], and Tennessee and Wisconsin (101 households, SAR 53%) [8] also report higher SARs. The former study was completed during a time of shelter-in-place policies. A retrospective study of 32 households of pediatric cases that relied on symptom ascertainment, also during a time of shelter-in-place, found a SAR of 46% [24]. Altogether, these studies have started to paint a picture of much higher secondary attack rates within households.

There are several likely explanations for why the SAR we report is the highest yet among US studies. Compared to previous studies, this study had longer follow-up, including weekly PCR testing to 21 days, combined with antibody testing at day 28. Longer follow-up is needed to capture potential tertiary cases (from sequential transmission) in the household. However, cases identified later during follow-up may also have been acquired in the community, as the study spanned seven months whilst the epidemic in North Carolina evolved from nursing homes, prisons, and meatpacking facilities; to frontline workers; to returning college students; and finally the general population. We suspect separately community-acquired cases are few amongst the household contacts in this study, but even limiting our SAR analysis to secondary cases detected within the first week of enrollment, the attack rate among household contacts is still >50%. Second, representation of racial and ethnic diversity has been limited in prior studies (>=70% white, non-Hispanic in each of the three aforementioned studies [7,8,23]). We found that risk factors for secondary infection in household contacts - including higher living density and obesity - were more frequent among households with participants who identified as non-white or Hispanic, who comprised 45% of our study sample. Third, although we excluded 22% of household contacts infected at baseline due to report of a common exposure as the index case, this proportion may in fact have been higher due to potential recall bias for common exposures. However, in our experience, a large proportion of these exposures still occur among family, if not the immediate household. In 44% of households, at least one household member (most often young children) declined to participate, which may have biased our estimate as well. Finally, the CO-HOST study was conducted during a time when the potentially more infectious 614G variant [25] predominated in North Carolina, involving >95% of our sample, paralleling its rise and dominance in the United States [18]. Overall, it is clear that SAR will vary in different settings and needs to be contextualized based on geography, risk groups, and the level of community transmission and public policies in effect at the time of the study.

Our data, with the majority of cases occurring within one week from illness onset in the index case, are consistent with previous modeling studies indicating that infectiousness peaks just before the onset of symptoms [3–5,26]. Practically speaking, this means that by the time the first case in the household is diagnosed, others are already incubating virus if not already testing positive. This is especially true when there are delays to testing or obtaining results, as was common in the first few months of the pandemic. Thus, public health messages to wear masks and self-isolate at onset of symptoms, while prudent, are unlikely to eliminate household spread, even if they were feasible in all households. Early and frequent testing, combined with agents for post-exposure prophylaxis, would be needed to substantially mitigate the impact of the virus on families that have been inoculated and not yet vaccinated [27]. Otherwise, mask wearing within a household at all times is preferable in households with unvaccinated members who are vulnerable to severe COVID-19.

The length of household quarantine is often problematic for COVID-positive persons and their households. Current recommendations worldwide favor a 14-day quarantine period for the entire household if one member is infected. However, compliance is difficult, especially for families with young children, those with limited resources, and those unable to work from home. If the quarantine period is decreased, the risk of onward transmission is increased, but the size of this risk remains an active subject of investigation [20,27]. One approach has been to reset the ‘quarantine clock’ for the entire household by 14 days each time a new household member is diagnosed, but this has further increased the burden and decreased compliance. In this study, two-thirds of household contacts were already infected at enrollment, a median of 6 days after symptom onset in the index case. We found that 94% of secondary cases were detected within 14 days from symptom onset of the index case, and resetting the clock on quarantine based on subsequent cases in the household was of incremental benefit (capturing an additional 4% of cases). This data supports the recommendation of a single 14-day quarantine for the entire household.

A novel finding of our study is the correlation of SARS-CoV-2 viral burden within households. Increased viral load increases infectivity *in vivo [25]*, and a recent study of 282 clusters in Spain (many involving household contacts) showed increased risk of transmission with shorter time to onset of symptoms among contacts as viral load increased [28]. Additionally, an increasing number of studies are confirming that greater viral burden (high viral load or lower Ct values by PCR) is associated with disease severity [21,29,30]. Now adding a third piece to this puzzle, we show that households seeded with a high viral load infection are more likely to have others with high viral loads, and therefore increased risk for severe illness. This implies that when a person is hospitalized, others in the same household may be at an even higher risk for a similar outcome compared to risk based on their individual risk factors (age, comorbidities) alone. Anecdotally, husbands and wives, siblings, and adult parents and children are not infrequently hospitalized in succession, though the prevalence of this is unknown. An inoculum effect may underlie this finding [31] and also explain why secondary cases in households appear to be overdispersed, with either most or all members infected, or none at all [6,32,33]. Viral load dynamics will no doubt continue to shape household transmission and the larger pandemic, as newer, potentially more infectious variants emerge even as vaccination decreases the “community viral load.”

To our knowledge, this is also the first study to show increased transmission in non-white US households. Though they experience similar rates of case fatality, African American/Black and Hispanic populations in the US experience disproportionately higher rates of SARS-CoV-2 infection and COVID-19–related mortality [34]. These racial disparities are thought to be due to differences in health care access and exposure risk that are driven by systemic societal inequities rather than individual biological or behavioral characteristics [35–38]. The CO-HOST study is consistent with this explanation. While the sample size was not sufficient to investigate drivers of the increased transmission in minority households, we found that high living density/household crowding, which was more common in the non-white households, was associated with increased transmission. Trends in home ownership, educational status, and living space within our data support the role of social vulnerabilities in modulating transmission risk within households, a major setting of SARS-CoV-2 transmission.

In our risk factors analysis, we found that spouses/partners and household members with obesity were at higher risk of becoming infected, while households of healthcare workers were less likely to become infected. All of the index cases in this study were symptomatic, hence we were unable to assess the likelihood of transmission from symptomatic versus asymptomatic cases. We were also unable to detect any impact of age or other comorbidities on acquisition of infection, likely due to the small effect size mediated through these variables and limited sample numbers. However, a meta-analysis has found that secondary attack rates are increased from symptomatic index cases in comparison to asymptomatic cases, adult index cases in comparison to child index cases, and in spouses compared to other family members [6].

In conclusion, SARS-CoV-2 transmits early and often among household members. While masking, physical distancing, and quarantining the whole household may reduce or prevent transmission beyond the household, these strategies are less effective and feasible within the household, especially in the setting of high viral load infections and crowded living spaces. Frequent point-of-care testing and prophylaxis in those at-risk for severe illness, and ultimately widespread and equitable distribution of vaccines, are needed to lessen the impact of COVID-19 within households and vulnerable communities.

## Supporting information

Strobe Check List

## Data Availability

Data is available on request for any interested researchers to allow replication of results provided all ethical requirements are met.

## ACKNOWLEDGEMENTS

We thank our wonderful CO-HOST study participants, Moby and the Chapel Hill CRS, and the UNC RDC team. Thanks to MIchelle Berrey, JoAnn Kuruc, and Dania Munson for help with protocol writing and submission; to Oksana Kharabora, Maureen Furlong, Amy James Loftis, Tia Belvin, and Dana Swilley for help with study preparation and implementation; to Joe Eron, Billy Fischer, and Ada Adimora for their input and support; and to Gabby Streeter for help with data analysis.

## SUPPORTING INFORMATION

**Figure S1.**
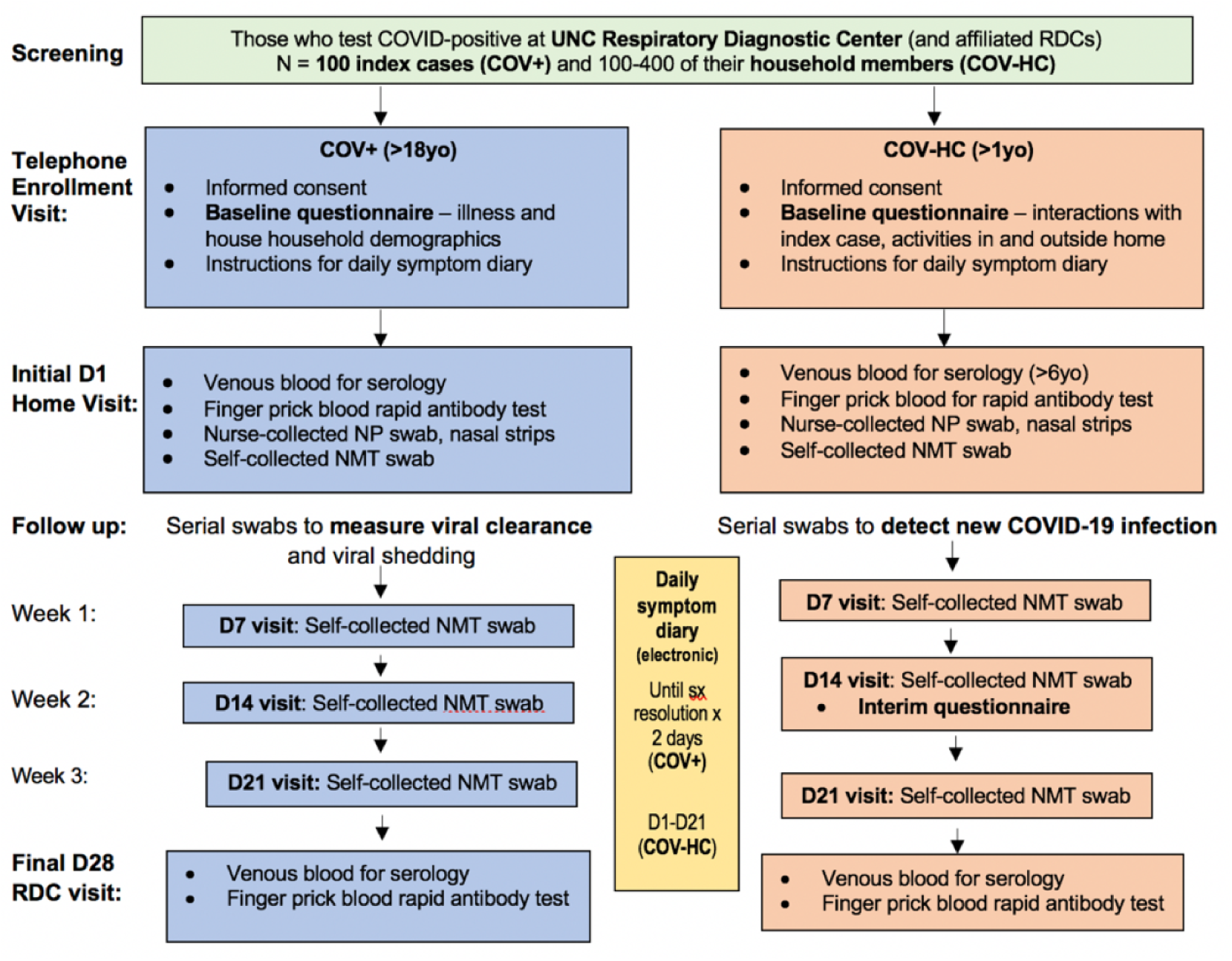
Schematic of CO-HOST Study Design.

**Table S1:**
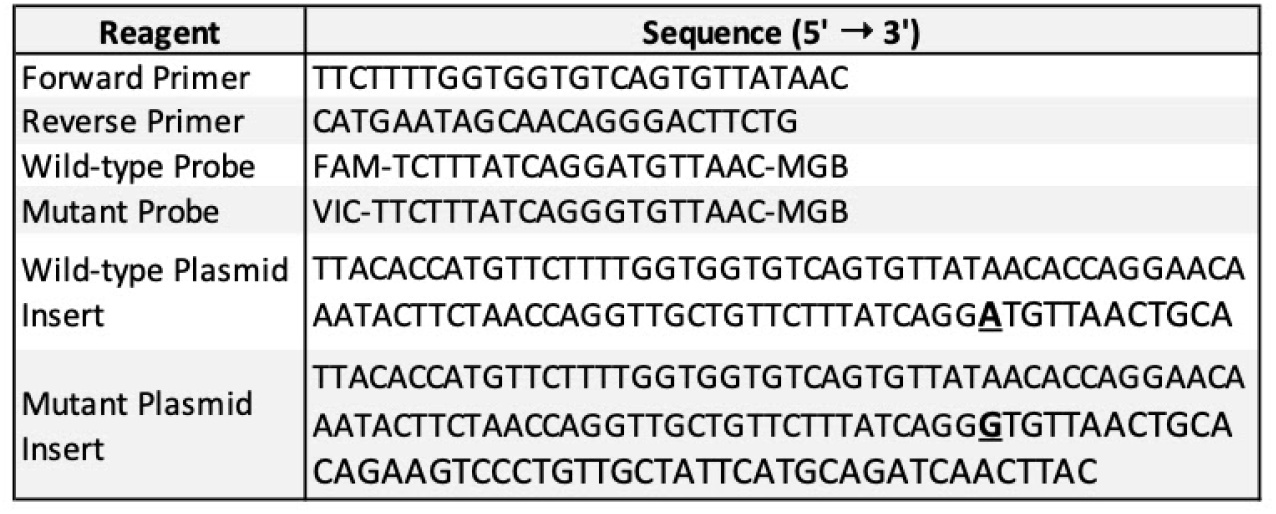
Sequences of primers, probes, and plasmids used for SARS-CoV-2 D614G genotyping by real-time PCR.

**Table S2.** Comorbidities of study participants.

| **INDIVIDUALS** | **Index (n)** | **Index (%)** | **HC (n)** | **HC (%)** |
| --- | --- | --- | --- | --- |
| *Underlying Conditions for Adults >18y\** | **92** | **%** | **134** | **%** |
| Cancer | 3 | 3.3 | 0 | 0.0 |
| Chronic lung disease | 1 | 1.1 | 2 | 1.5 |
| Asthma | 9 | 9.8 | 19 | 14.2 |
| Daily smoker | 2 | 2.2 | 11 | 8.2 |
| Diabetes | 6 | 6.5 | 12 | 9.0 |
| High blood pressure | 16 | 17.4 | 30 | 22.4 |
| Heart disease | 2 | 2.2 | 2 | 1.5 |
| Chronic kidney disease | 1 | 1.1 | 0 | 0.0 |
| Recent (within past 2 weeks) or current pregnancy | 3 | 3.3 | 5 | 3.7 |
| BMI >30 | 35 | 38.0 | 43 | 32.1 |
| BMI 25-29.9 | 24 | 26.1 | 37 | 27.6 |
| *Underlying Conditions for Adults >50y\** | **23** | **%** | **42** | **%** |
| Asthma | 2 | 8.7 | 2 | 4.8 |
| Daily smoker | 0 | 0.0 | 5 | 11.9 |
| Diabetes | 5 | 21.7 | 7 | 16.7 |
| High blood pressure | 6 | 26.1 | 21 | 50.0 |
| BMI >30 | 11 | 47.8 | 14 | 33.3 |
| BMI 25-29.9 | 5 | 21.7 | 16 | 38.1 |
| BMI >30 and one or more co-morbidity (adults >18y) (n = 92, 134) | 22 | 23.9 | 25 | 18.7 |
| BMI >30 and one or more co-morbidity (adults >50y) (n = 23, 42) | 8 | 34.8 | 12 | 28.6 |
| *\*No adults >18y with HIV or chronic liver disease* |  |  |  |  |

**Table S3.** Household demographics.

| **HOUSEHOLDS** | **(n=100)** | **(%)** |
| --- | --- | --- |
| *Household Size* |  |  |
| 2 People | 27 | 27.0 |
| 3 People | 23 | 23.0 |
| 4 People | 22 | 22.0 |
| 5 or more people | 28 | 28.0 |
| *Home Ownership (n = 97)* |  |  |
| Single-family home/townhome occupied by owner | 61 | 62.9 |
| Single-family home/townhome occupied by renter | 25 | 25.8 |
| Apartment occupied by renter | 10 | 10.3 |
| Other | 1 | 1.0 |
| *Rooms in the House\** |  |  |
| 2 or fewer rooms | 10 | 10.0 |
| 3-5 rooms | 43 | 43.0 |
| 6 or more rooms | 47 | 47.0 |
| *\*including bedrooms, kitchen, and common rooms, but not bathrooms or garage* |  |  |
| *Living Space* |  |  |
| <500 sq feet (<46.5 sq m) | 3 | 3.0 |
| 500-1000 sq feet (46-93 sq m) | 17 | 17.0 |
| 1000-2000 sq feet (93-186 sq m) | 33 | 33.0 |
| >2000 sq feet (>186 sq m) | 42 | 42.0 |
| Unknown | 5 | 5.0 |

