## Supplementary material for "High household transmission of SARS-CoV-2 in the United States: living density, viral load, and disproportionate impact on communities of color": Strobe Check List

STROBE Statement—checklist of items that should be included in reports of observational studies

| | Item No | Recommendation |
|---|---|---|
| **Title and abstract** | 1 | (*a*) Indicate the study's design with a commonly used term in the title or the abstract<br>**Done. Page 1, line 17**<br>*"This is a prospective observational study."* |
| | | (*b*) Provide in the abstract an informative and balanced summary of what was done and what was found<br>**Done. Page 1, lines 14-40.** |
| **Introduction** | | |
| Background/rationale | 2 | Explain the scientific background and rationale for the investigation being reported<br>**Done. Page 3, lines 71-91. Introduction** |
| Objectives | 3 | State specific objectives, including any prespecified hypotheses.<br>**Done. Page 6, line 192-199 "Study Objectives and Outcomes".**<br>*"The primary objective was to evaluate the secondary household attack rate among household members of persons quarantined in their home after testing positive for SARS-CoV-2.*<br>*The primary study endpoint was SARS-CoV-2 infection in the household contacts as determined by real-time PCR of nasopharyngeal or nasal swabs for SARS-CoV-2 at any of the timepoints or evidence of seroconversion during the study based on anti-SARS-CoV-2 antibody testing.*<br>*A secondary objective was to assess individual and household risk factors associated with SARS-CoV-2 transmission in the household."* |
| **Methods** | | |
| Study design | 4 | Present key elements of study design early in the paper<br>**Done. Page 4, lines 117-148. "Recruitment, screening and enrolment" and Figure S1 Schematic of CO-HOST study design** |
| Setting | 5 | Describe the setting, locations, and relevant dates, including periods of recruitment, exposure, follow-up, and data collection<br>**Done. Page 4, lines 113-116 "Study setting"**<br>*"Index cases were recruited after testing at the Respiratory Diagnostic Center at the University of North Carolina School of Medicine [10]. Participants were visited between 3-4 times at their private homes using a mobile unit van and returned to the Respiratory Diagnostic Center for the final study visit."* |
| Participants | 6 | *Cohort study*—Give the eligibility criteria, and the sources and methods of selection of participants. Describe methods of follow-up<br>**Done. Page 4, lines 117-124. "Recruitment, screening and enrolment"**<br>*"Inclusion criteria for the index cases included any patient 18 years of age or older with a positive qualitative nasopharyngeal (NP) swab for SARS-CoV-2 obtained at UNC Hospitals, willingness to self-isolate at home for a 14-day period, willingness to participate in all required study activities for the entire 28-day duration of the study, living with at least one household contact who was also willing to consent to study follow-up, and living within reasonable driving distance (<1 hour) suitable for home visits by the study team. Inclusion criteria for household contacts of index patients included age greater than 1 year, and currently living in the same home as the index case without plans to leave to live elsewhere through the end of the 28-day study."*<br><br>*Case-control study*—Give the eligibility criteria, and the sources and methods of case ascertainment and control selection. Give the rationale for the choice of cases and controls<br>*Cross-sectional study*—Give the eligibility criteria, and the sources and methods of selection of participants |
| | | (*b*) *Cohort study*—For matched studies, give matching criteria and number of exposed |

| | | |
|---|---|---|
| | | and unexposed.<br>*Case-control study*—For matched studies, give matching criteria and the number of controls per case |
| Variables | 7 | Clearly define all outcomes, exposures, predictors, potential confounders, and effect modifiers. Give diagnostic criteria, if applicable<br>**Done. Page 6, line 195-197 "Study Objectives and Outcomes".**<br>*The primary study endpoint was SARS-CoV-2 infection in the household contacts as determined by real-time PCR of nasopharyngeal or nasal swabs for SARS-CoV-2 at any of the timepoints or evidence of seroconversion during the study based on anti-SARS-CoV-2 antibody testing.* |
| Data sources/ measurement | 8* | For each variable of interest, give sources of data and details of methods of assessment (measurement). Describe comparability of assessment methods if there is more than one group.<br>**Done. Page 6, line 200-207 " Data entry, handling, storage and security"**<br>*"After giving written consent, the participants were given a study identification number, which was used in all future datasets for participant anonymity. Collected data were entered in real-time using electronic Case Report Forms (eCRF) developed on a REDCap (Research Electronic Data Capture) database. Any data collected on paper format was entered by a study staff member and then checked by the study coordinator. Daily symptom diaries were entered directly into the REDCap database by the participants and were checked by study staff for completion and inconsistencies. Laboratory related data were extracted directly from laboratory equipment and uploaded to the database. The study was conducted in compliance with Good Clinical Practice."* |
| Bias | 9 | Describe any efforts to address potential sources of bias.<br>**Done. Page 6, lines 208-211**<br>*"For each household, if multiple participants were SARS-CoV-2 positive at enrollment, we defined the index case as the person with the earliest onset of infection based on onset of symptoms and known date(s) of PCR test positivity. If this was ambiguous and to prevent bias, then baseline antibody positivity was also used as evidence of less recent infection."* |
| Study size | 10 | Explain how the study size was arrived at<br>**Done. Page 6, lines 189-191, "Sample size determination"**<br>*"This is a prospective observational study. The planned target enrollment was 200 households. The study was stopped prior to reaching this target due to funding considerations."* |
| Quantitative variables | 11 | Explain how quantitative variables were handled in the analyses. If applicable, describe which groupings were chosen and why<br>**Done. Page 7, lines 237-244, "Statistical analysis"**<br>*"We determined whether nasopharyngeal SARS-CoV-2 viral loads were correlated within households (whether persons in the same household were more likely to have similar NP viral loads) by the intraclass correlation coefficient (ICC), which compares within versus between households variation of baseline NP viral loads. For those participants who did not complete an NP swab on study day 1, we used a transformed NMT viral load to impute the missing NP value. The transformation formula was derived from a linear regression equation generated from >100 study participants with positive viral load from both NP and NMT swabs on study day 1 [12]. To determine whether NP viral load in index cases was associated with secondary cases in the same household, we dichotomized the NP viral load with a cutoff of* |

| | | |
|---|---|---|
| | | *1x10^6 viral copies/ul and compared the proportion of transmission events."* |
| Statistical methods | 12 | (*a*) Describe all statistical methods, including those used to control for confounding<br>**Done. Pages 6-7, lines 208-254.** |
| | | (*b*) Describe any methods used to examine subgroups and interactions.<br>**N/A** |
| | | (*c*) Explain how missing data were addressed<br>**Done. Page 7, lines 223-226 "Statistical analysis"**<br>*"Household contacts were excluded from the SAR analysis if they missed all follow-up study visits (n=6) or were symptomatic with negative PCR testing but missing antibody data at day 28 (n=1). Among those included in the analysis, the rate of missing data was low (<5%); thus, we did not impute missing data."* |
| | | (*d*) *Cohort study*—If applicable, explain how loss to follow-up was addressed<br>**N/A**<br>*Case-control study*—If applicable, explain how matching of cases and controls was addressed<br>*Cross-sectional study*—If applicable, describe analytical methods taking account of sampling strategy |
| | | (*e*) Describe any sensitivity analyses<br>**Done. Page 7, lines 229-233. "Statistical analysis"**<br>*"In the primary SAR analysis, all secondary cases were presumed due to household transmission (not community-acquired). Sensitivity analyses were performed excluding secondary cases already infected at baseline or excluding secondary cases identified at day 14 or later that may have been acquired outside the household. The SAR for households was calculated as the proportion of households with at least one secondary case identified in the household during the 28-day follow-up."* |

**Results**

| | | |
|---|---|---|
| Participants | 13* | (a) Report numbers of individuals at each stage of study—eg numbers potentially eligible, examined for eligibility, confirmed eligible, included in the study, completing follow-up, and analysed<br>**Done. Page 10, Figure 2** |
| | | (b) Give reasons for non-participation at each stage<br>**N/A** |
| | | (c) Consider use of a flow diagram<br>**Done. Page 10, Figure 2** |
| Descriptive data | 14* | (a) Give characteristics of study participants (eg demographic, clinical, social) and information on exposures and potential confounders<br>**Done. Page 9, Table 1: Demographics of Study Participants** |
| | | (b) Indicate number of participants with missing data for each variable of interest<br>**Done. Page 9, Table 1: Demographics of Study Participants.** |
| | | © *Cohort study*—Summarise follow-up time (eg, average and total amount)<br>**Done. Page 9, line 301-302.**<br>*"Of 100 households with 304 study participants (100 index cases and 204 HCs) included in the analysis, 99 households completed one month follow-up."* |
| Outcome data | 15* | *Cohort study*—Report numbers of outcome events or summary measures over time<br>**Done. Page 10 Figure 2 and Page 11, Figure 3.** |
| | | *Case-control study*—Report numbers in each exposure category, or summary measures of exposure |

| | | |
|---|---|---|
| | | *Cross-sectional study*—Report numbers of outcome events or summary measures |
| Main results | 16 | (*a*) Give unadjusted estimates and, if applicable, confounder-adjusted estimates and their precision (eg, 95% confidence interval). Make clear which confounders were adjusted for and why they were included<br>**Done. Page 10 Figure 2 and Page 11, Figure 3.** |
| | | (*b*) Report category boundaries when continuous variables were categorized<br>**N/A** |
| | | (*c*) If relevant, consider translating estimates of relative risk into absolute risk for a meaningful time period<br>**N/A** |
| Other analyses | 17 | Report other analyses done—eg analyses of subgroups and interactions, and sensitivity analyses.<br>**Done. Tables 1-5.** |
| **Discussion** | | |
| Key results | 18 | Summarise key results with reference to study objectives<br>**Done. Page 15, lines 422-431.**<br>*"Household transmission is one of the main drivers of the SARS-CoV-2 pandemic. By incorporating timely recruitment of index cases, prospective sampling to 21 days regardless of symptom status, and diverse representation, we show that household transmission occurs in the majority of COVID-positive North Carolina households. The overall secondary attack rate in our sample was 60%, rising to 70% in minority households and 91% in households with higher living density. Importantly, we show not only that those infected with a high viral load are more likely to transmit virus to other members of the household, but that they seed other high-viral load infections, putting the entire household at higher risk for more severe illness [21]. Spread within the household happens quickly, often with one or more household members already infected by the time the first case in the household is diagnosed."* |
| Limitations | 19 | Discuss limitations of the study, taking into account sources of potential bias or imprecision. Discuss both direction and magnitude of any potential bias.<br>**Done.**<br>**Page 15, lines 447-452**<br>*"However, cases identified later during follow-up may also have been acquired in the community, as the study spanned seven months whilst the epidemic in North Carolina evolved from nursing homes, prisons, and meatpacking facilities; to frontline workers; to returning college students; and finally the general population. We suspect separately community-acquired cases are few amongst the household contacts in this study, but even limiting our SAR analysis to secondary cases detected within the first week of enrollment, the attack rate among household contacts is still >50%."*<br>***Page 15, lines 456-460***<br>*"….although we excluded 22% of household contacts infected at baseline due to report of a common exposure as the index case, this proportion may in fact have been higher due to potential recall bias for common exposures. However, in our experience, a large proportion of these exposures still occur among family, if not the immediate household. In 44% of households, at least one household member (most often young children) declined to participate, which may have biased our estimate as well."*<br>**Page 17, lines 505-507**<br>*"While the sample size was not sufficient to investigate drivers of the increased transmission in minority households, we found that high living density/household crowding, which was more common in the non-white households, was associated with increased transmission."*<br>**Page 17, lines 511-515** |

| | | |
|---|---|---|
| | | *"All of the index cases in this study were symptomatic, hence we were unable to assess the likelihood of transmission from symptomatic versus asymptomatic cases. We were also unable to detect any impact of age or other comorbidities on acquisition of infection, likely due to the small effect size mediated through these variables and limited sample numbers."* |
| Interpretation | 20 | Give a cautious overall interpretation of results considering objectives, limitations, multiplicity of analyses, results from similar studies, and other relevant evidence<br>**Done. Page 17 lines 519-524 "Discussion"**<br>*"In conclusion, SARS-CoV-2 transmits early and often among household members. While masking, physical distancing, and quarantining the whole household may reduce or prevent transmission beyond the household, these strategies are less effective and feasible within the household, especially in the setting of high viral load infections and crowded living spaces. Frequent point-of-care testing and prophylaxis in those at-risk for severe illness, and ultimately widespread and equitable distribution of vaccines, are needed to lessen the impact of COVID-19 within households and vulnerable communities."* |
| Generalisability | 21 | Discuss the generalisability (external validity) of the study results<br>**Done. Page 15-17 lines 423-524, throughout the "Discussion"** |
| **Other information** | | |
| Funding | 22 | Give the source of funding and the role of the funders for the present study and, if applicable, for the original study on which the present article is based<br><br>**Done. Page 17, lines 519-524 "FUNDING"**<br>*"Research was supported by funds and charitable contributions from the UNC Department of Medicine, UNC COVID-19 Response Fund/Health Foundation, a Gillings Innovations Lab Award, and the National Center for Advancing Translational Sciences (NCATS), National Institutes of Health, through Grant Award Number UL1TR002489. Rapid antibody tests were provided by Biomedomics Inc, Morrisville, NC."* |

*Give information separately for cases and controls in case-control studies and, if applicable, for exposed and unexposed groups in cohort and cross-sectional studies.

**Note:** An Explanation and Elaboration article discusses each checklist item and gives methodological background and published examples of transparent reporting. The STROBE checklist is best used in conjunction with this article (freely available on the Web sites of PLoS Medicine at http://www.plosmedicine.org/, Annals of Internal Medicine at http://www.annals.org/, and Epidemiology at http://www.epidem.com/). Information on the STROBE Initiative is available at www.strobe-statement.org.
